# The Impact of Influenza Vaccination on Antibiotic Prescribing in Older Adults: A Self-Controlled Case Series Analysis

**DOI:** 10.64898/2025.12.02.25341465

**Authors:** Thomas Spain, Marc Y Henrion, David Singleton, Roberto Vivancos, Valerie Decraene, Daniel Hungerford, Neil French

## Abstract

**Background:** Influenza imposes a significant health burden on older adults, with influenza-like illness (ILI) a key driver of antibiotic prescribing, contributing to unnecessary antibiotic use and antimicrobial resistance (AMR). Influenza vaccination may reduce antibiotic prescribing; however, robust evidence remains limited. To address this, we examined the association between influenza vaccination and antibiotic prescribing in individuals aged 65 and older in England, using a self-controlled case series (SCCS) approach.

**Methods:** We conducted a SCCS analysis using electronic primary care health records from the Clinical Practice Research Datalink (CPRD), covering eight influenza seasons (2011–2019). Eligible individuals had at least one influenza season in which they were both vaccinated and unvaccinated. The primary outcome was days prescribed antibiotics, focusing on respiratory tract infection (RTI)-linked prescribing. To account for seasonal variation in influenza circulation, we initially examined antibiotic prescribing from September-April and further conducted a post-hoc analysis restricted to January-April to assess the vaccine’s effect during peak influenza activity. Incidence rate ratios (IRRs) were estimated using conditional Poisson regression models, adjusting for comorbidities, healthcare interactions, and other vaccinations.

**Results:** The analysed extract included 268,352 individuals, of whom 48,329 met the inclusion criteria. On average, individuals were prescribed antibiotics for 8 days per influenza season. Findings suggest influenza vaccination may reduce overall and RTI-related antibiotic prescribing, particularly during peak influenza season. The IRR for RTI-linked prescribing decreased from 0.97 (95% CI: 0.95–0.98) for September-April to 0.91 (95% CI: 0.89–0.93) for January-April.

**Conclusion:** Influenza vaccination contributes to reduced RTI-related antibiotic prescribing in older adults during the influenza season.

**Key points:** There is limited robust evidence on the impact of influenza vaccines on antibiotic prescribing. To mitigate bias, we used electronic medical records in a self-controlled case series, showing modest reductions in respiratory tract infection-related prescribing in influenza vaccinated older adults.

## Introduction

Influenza imposes a significant burden on older adults, with this population experiencing the highest rates of complications, hospitalizations, and mortality associated with infection (1–3). Influenza-like illness is associated with increased antibiotic prescribing, particularly during periods of heightened flu activity (4), contributing to unnecessary antibiotic use and potentially the broader public health challenge of antimicrobial resistance (AMR). Respiratory tract infections (RTIs) account for nearly half of antibiotic consultations in primary care, and the majority of antibiotic prescribing in England occurs in primary care settings (5, 6).

Vaccines aimed at preventing RTIs are therefore particularly relevant to reducing antibiotic prescribing (7). Vaccination has the potential to reduce antibiotic prescribing either by reducing the frequency and/or severity of influenza like illness attendances in primary care.

Influenza vaccination remains a critical public health measure to prevent illness and mitigate severe outcomes. Bacterial coinfections are clinically well-recognised as associated with influenza-attributed deaths, with rates reported as notably higher during pandemic period rather than in a typical season of influenza. For example, up to 34% of 2009 pandemic influenza A virus (H1N1) infections have bacterial coinfections (8, 9). This risk is the major driver of prescribing in primary care. Vaccination should reduce antibiotic-requiring secondary bacterial respiratory infections, especially in young children and the elderly (10, 11).

In the UK, inactivated influenza vaccination is freely available to people over 65 years, with uptake reported consistently over 70% (72% in 2018/19) (12). The estimated reduction in medically-attended influenza infection associated with vaccination varies season to season, but was estimated at 49% in 2018/19 against medically attended influenza infection in individuals aged 65 years and over (13). Given that seasonal influenza vaccines are widely used among older and other at-risk populations in many countries (14), understanding their impact on antibiotic use is of substantial policy relevance. However, evidence for the impact of influenza vaccination on antibiotic prescribing remains limited. A systematic review highlighted a lack of robust studies for evidence of vaccine impact on antibiotic prescribing (15), highlighting a single randomized controlled trial (RCT) that suggested a reduction in antibiotic prescribing associated with a live attenuated influenza vaccination in healthy, low risk adults, which is not currently used in the UK (16). Observational studies have faced challenges with critical risks of bias, particularly confounding by indication, with a limited number of studies with a low risk of bias producing heterogeneous results (17). Larger studies employing robust analytical methods are necessary to accurately evaluate the impact of vaccination on antibiotic prescribing. To address these challenges, our study applies the self-controlled case series (SCCS) design, which controls for time-varying individual characteristics and is particularly well-suited for evaluating effects of seasonal vaccines.

This study examines the effect of seasonal influenza vaccination on antibiotic prescribing among individuals aged 65 years and older in England attending primary care. The analysis spans eight influenza seasons (September 2011 to August 2019) and employs the self-controlled case series (SCCS) methodology to minimise confounding (18).

## Methods

### Data source

Electronic medical record (EMR) data extracted from the Clinical Practice Research Datalink (CPRD) Aurum forms the primary dataset for this study. CPRD Aurum, contains routinely collected EMR data from primary care practices in the UK using EMIS Web practice management software. These practices represent approximately 13% of the primary care-registered population of England and are broadly representative of the general population (19). The dataset includes GP-entered codes capturing symptoms, diagnoses, diagnostic tests, comorbidities, prescriptions, and other clinical information. CPRD Aurum data is structured across multiple interrelated tables, enabling comprehensive measurement of exposures, outcomes, and covariates relevant to this study.

### Study period, population and exposure

The study period spanned eight full influenza surveillance seasons, ranging from 1 September 2011 to 31 August 2019. Elderly (65 or older) persons were eligible to be vaccinated from 1 September each year. A full influenza season was defined as 1 September to 31 August in the following calendar year.

People were considered eligible for inclusion if they were registered with a GP prior to 1 September 2011 and did not transfer in or out of practice between 1 September 2011 and 31 August 2019. People who died during this study period were eligible, but the full influenza season in which they died was not included in analysis. This is because deteriorating health status during a season could independently influence both the decision to vaccinate, timing of vaccination and/or antibiotic prescribing within that season, introducing within-person confounding.

The primary exposure was influenza vaccination, defined as a non-time-varying binary variable denoting a record of an influenza vaccination during its corresponding full influenza surveillance season. The list of CPRD Aurum codes used to classify the exposure variable, the outcome variables, comorbidities and other explanatory variables are available on Github (https://github.com/tomjspain/IVARflu).

As the study used an SCCS design, people who were vaccinated in every influenza season and people who were unvaccinated in every influenza season during the study period were excluded, in order for a comparison between vaccinated and unvaccinated seasons to be possible within each individual.

### Outcome

The primary outcome for this study was number of days prescribed for any antibiotic between the first day of September each season until the last day of April the following year. This period was selected a-priori to match the start of influenza vaccine offer in England and the average end of seasonal influenza season in the England. We also conducted post-hoc analyses to more accurately reflect periods of high influenza circulation while excluding early-season variability in vaccination timing, shifting the start month from September to January, whilst maintaining the last day of April as the end date. The number of days for all prescriptions were derived from their original issues, but where this information was missing, the median number of days for the type of antibiotic (such as amoxicillin) was imputed. The secondary outcome for the study was the number of prescriptions within decreasing seasonal timeframes, as described above.

These outcomes measures were measured for all primary care attendances and for two condition subgroups. The first subgroup was respiratory tract infection (RTI)-linked antibiotic prescriptions within decreasing seasonal timeframes, where RTI-linked antibiotics were defined as any antibiotics from a prescription which was issued within seven days of an RTI observation. Urinary tract infections (UTIs) were selected as a control condition where we hypothesised that UTI rates would be unaffected by influenza vaccination. UTI-linked antibiotics were defined as any antibiotic from a prescription which was issued within seven days of a UTI observation.

A seven-day attribution window was chosen to accommodate delayed or “back-up” prescribing, a common UK primary care strategy in which a prescription is issued but the patient is advised to delay collection or initiation pending symptom progression.

Observational data indicate that the advised delay period is typically around three days but can extend to a week in some settings (20),so a seven-day window was considered a pragmatic upper bound that would capture the great majority of genuinely condition-linked prescriptions without materially extending misattribution to unrelated, later illness episodes.

Antibiotics were not restricted to specific drug classes for either subgroup; rather, any antibiotic prescription issued within the specified window of a relevant diagnostic observation (RTI or UTI) was attributed to that condition.

### Comorbidities

A patient is defined as having a comorbidity if code matching the condition is observed at least once per patient-influenza season. Scores associated with these were summed to provide a combined comorbidity score per patient-influenza season. Where no comorbidity is observed in an influenza season the patient’s score for that season was anchored to their score from their first season of follow-up, calculated as the higher of the two values; this was applied because most comorbidities considered are chronic and not expected to resolve over time, such that an apparent decrease in score between seasons more plausibly reflect under-recording than genuine improvement. The combined score was utilised as described by Gagne et al (21). Specifically, the condition list and corresponding weights presented in Table 3 of Gagne et al (21) were used to derive the combined score.

### Statistical analysis

The Self-Controlled Case Series (SCCS) design compares outcome rates within individuals across exposed and unexposed periods, thereby controlling by design for all fixed individual characteristics (e.g., sex, deprivation, underlying morbidity) (18). In this study, exposed time was defined as the seasonal timeframe in which an individual received influenza vaccination, and unexposed time as the corresponding window in seasons without vaccination. Incidence rate ratios (IRRs) thus estimate the relative rate of antibiotic-prescribing days during vaccinated versus unvaccinated seasons within the same person. The SCCS approach assumes that outcome occurrence does not influence subsequent vaccination within the annual seasonal observation window and that follow-up is not truncated by the outcome; these considerations informed our choice of seasonal risk windows and covariate adjustment.

SCCS models were fitted using a conditional Poisson regression model. Conditional Poisson regression models were fitted using the “gnm” package in R. Ninety five percent confidence intervals for model covariates were calculated using the Wald method, based on the standard errors of the estimated coefficients. This approach was used for the primary multivariable model and all sensitivity analyses.

The incidence rate ratio (IRR) represents the relative rate of days prescribed antibiotics in a vaccinated seasonal timeframe compared to its corresponding unvaccinated seasonal timeframe, estimated using the fully adjusted SCCS model. The model included full influenza season as a categorical variable, while influenza vaccine, and herpes zoster vaccine (live shingles vaccine) were treated as binary indicators. Pneumococcal vaccine (single dose 23 valent pneumococcal polysaccharide vaccine) was used, as this is the formulation offered to adults aged 65 and over under the UK’s national immunisation schedule; the conjugate vaccine (PCV13) was not routinely offered to this age group outside of specific clinical high-risk groups and so was not considered. Vaccination status for each of these covariates was updated once per influenza season, consistent with the comorbidity score described above, such that an individual’s status could change between seasons if vaccinated during the study period. Pneumococcal vaccine is offered to 65+ year olds and was included because of the potential associated effect on bacterial respiratory infections. Since September 2013 live shingles vaccine has been offered to 70+ year olds and was included as a measure of healthcare use and healthcare seeking behaviour in relation to vaccines (22). Number of GP interactions, combined sum comorbidity score and the number of days prescribed any antibiotic during the May-August period preceding each influenza sfeason were included as continuous covariates to accommodate time-varying baseline risk. Additionally, an interaction term between age in season (continuous, centred at 65) and influenza vaccine in season (binary) was included. This interaction allowed the relationship between age and prescribing to differ between vaccinated and unvaccinated seasons, including for individual who were unvaccinated in only one or a small number of seasons; in such cases, the age-specific effect within their unvaccinated season was accounted for via this term rather than treated as equivalent to an unvaccinated season occurring at a different age. The model incorporated an offset term, defined as the natural logarithm of the number of days in the seasonal timeframe, to account for varying observation periods.

### Sensitivity analyses

Several sensitivity analyses were conducted to assess the robustness of the findings. These included models excluding the herpes zoster vaccination covariate and unadjusted models including only the influenza vaccination indicator. Additionally, all analyses were repeated across progressively narrower seasonal time windows to examine the association during periods of peak influenza circulation.

To assess residual confounding, off-season antibiotic prescribing (May-August of the prior year) was modelled as a negative control outcome (23). We additionally tested whether prior-season prescribing predicted subsequent vaccination uptake within individuals, as a further check on reverse-causation bias (see Limitations for full results). We also assessed sensitivity to vaccination timing by restricting the vaccinated period to at least 14 days post-vaccination and, separately, by excluding individuals vaccinated after 30 November, reclassifying affected patient-seasons as unvaccinated and reapplying the partially-vaccinated inclusion criteria in each case. Finally, as prescribing durations were imputed for a small number of prescriptions, the September-April models were repeated on a complete-case analysis sample excluding patient-seasons for which duration had been imputed, again reapplying the partially-vaccinated inclusion criteria to the resulting sample. UTI-related prescribing was additionally stratified by sex, given known difference in UTI incidence between men and women.

### Ethics

All data in this study were anonymised. CPRD’s Research Data Governance (RDG) granted approval (reference 21_000457). Rolling ethical approval is obtained for CPRD from the Health Research Authority (reference number 05/MRE04/87).

## Results

From an initial extract of 268,352 individuals, 48,329 met the inclusion criteria for the SCCS analysis, as 136,752 were vaccinated every year, 41,927 never received a vaccine, and a further 23,336 individuals were excluded after no longer meeting the partially vaccinated criteria following excluding death in season, and those with missing off-season outcomes preceding their first season of analysis (Figure 1). Demographic characteristics of the included participants are shown in Table 1.

**Figure 1:**
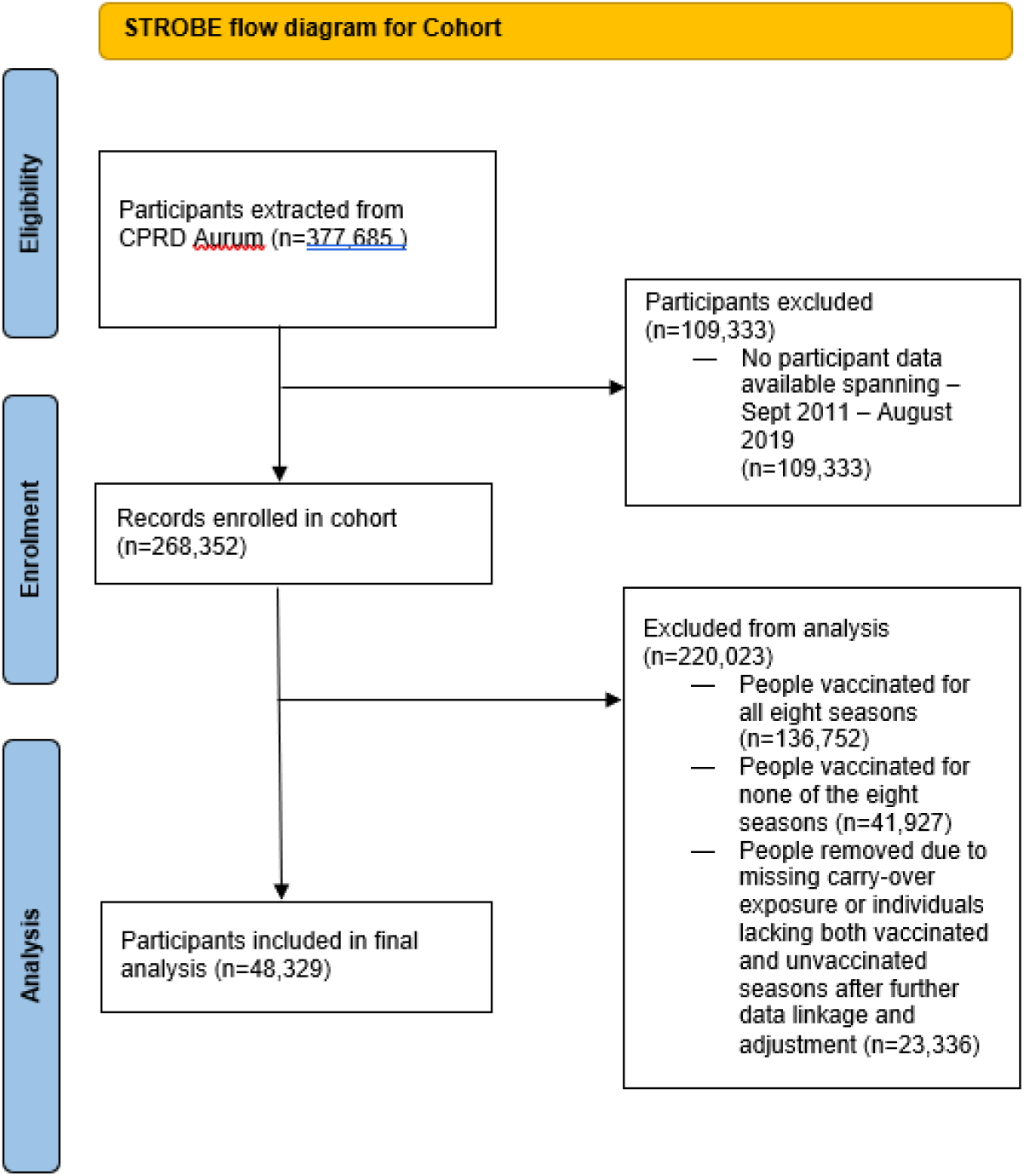
PRISMA flow diagram of final analysed cohort

**Table 1:** Characteristics of the final analysed cohort (n=48,329) meeting inclusion criteria.. Percentage or Median (IQR) reported.

| Characteristic | Percentage or Median [IQR] |
| --- | --- |
| **Gender (female)** | 57.9% |
| **IMD quintile** |  |
| 1 (most deprived) | 14.8% |
| 2 | 17.6% |
| 3 | 20.0% |
| 4 | 22.9% |
| 5 (least deprived) | 24.5% |
| Missing | 0.1% |
| **Practice region** |  |
| East Midlands | 1.5% |
| East of England | 5.7% |
| London | 14.3% |
| North East | 4.5% |
| North West | 16.7% |
| South Central | 21.3% |
| South West | 11.4% |
| West Midlands | 18.9% |
| Yorkshire and the Humber | 3.8% |
| Missing | 1.9% |
| **Urban-rural status** |  |
| Rural | 20.6% |
| Urban | 79.3% |
| Missing | 0.1% |
| **Median number of seasons of follow-up** | 8 [6, 8] |
| **Median age in season** | 76.5 [71.5, 82.5] |
| **Median patient-clinical staff interactions in season** | 7.0 [4.0, 11.5] |
| **Median combined sum comorbidity score** | 0 [0, 1] |

### Demographic Characteristics

Table 1 provides an overview of the cohort’s demographic and clinical characteristics. The median age during each influenza season was 76.5 years (IQR: 71.5–82.5), and the cohort consisted of 57.9% females. Most participants resided in urban areas (79.3%) and were predominantly in the two least deprived quintiles of socio-economic deprivation (47.4%). The median number of patient-clinical staff interactions per season was 7.0 (IQR: 4.0–11.5).

Seasonal variation in the number of individuals contributing to each influenza season and median age are shown in Supplementary Figure S1.

### Seasonal trends in antibiotic prescribing and influenza vaccinations

A progressive decrease in antibiotic prescribing rates was observed over the eight study seasons (Figure 2), vaccination coverage fluctuated during the study period and ranged between 58 and 69 percent.

**Figure 2:**
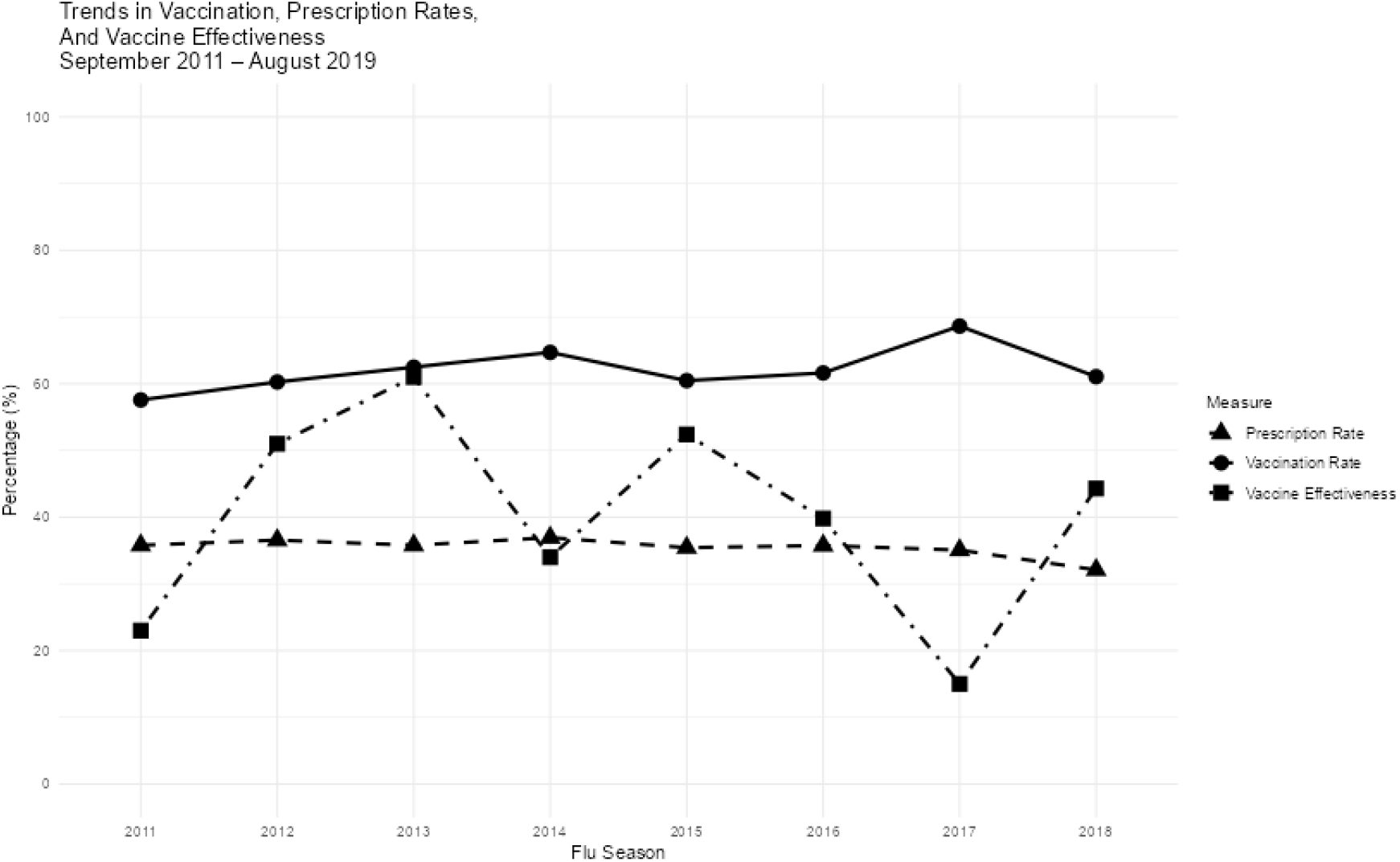
Seasonal vaccination and antibiotic prescription rates (at least one prescription within a season) in the elderly population (those > 65 years of age) in England.

### Antibiotic prescribing

In seasons when participants received influenza vaccine there was evidence for decreased prescribing of antibiotics used for the management of RTIs, indicated by an IRR of 0.97 (95% CI: 0.95–0.98). However, there was evidence of an excess of prescribing of all antibiotics, with an IRR of 1.03 (95% CI: 1.02–1.03) and a substantial excess of prescribing for UTI, demonstrated with an IRR of 1.15 (95% CI: 1.12-1.18).

RTI-associated prescribing shows a marked decrease from September-April, dropping substantially to 0.91 (95% CI: 0.89–0.93) for January-April (Figure 3), indicating a reduction in prescribing during vaccinated periods as the timeframe narrows to peak influenza season. For all prescribing, the IRR is unchanged (IRR=1.02, 95% CI: 1.01–1.02) for January-April. UTI-associated prescribing remains elevated during peak influenza season, going to an IRR of 1.15 (95% CI: 1.11–1.19) for January-April.

**Figure 3:**
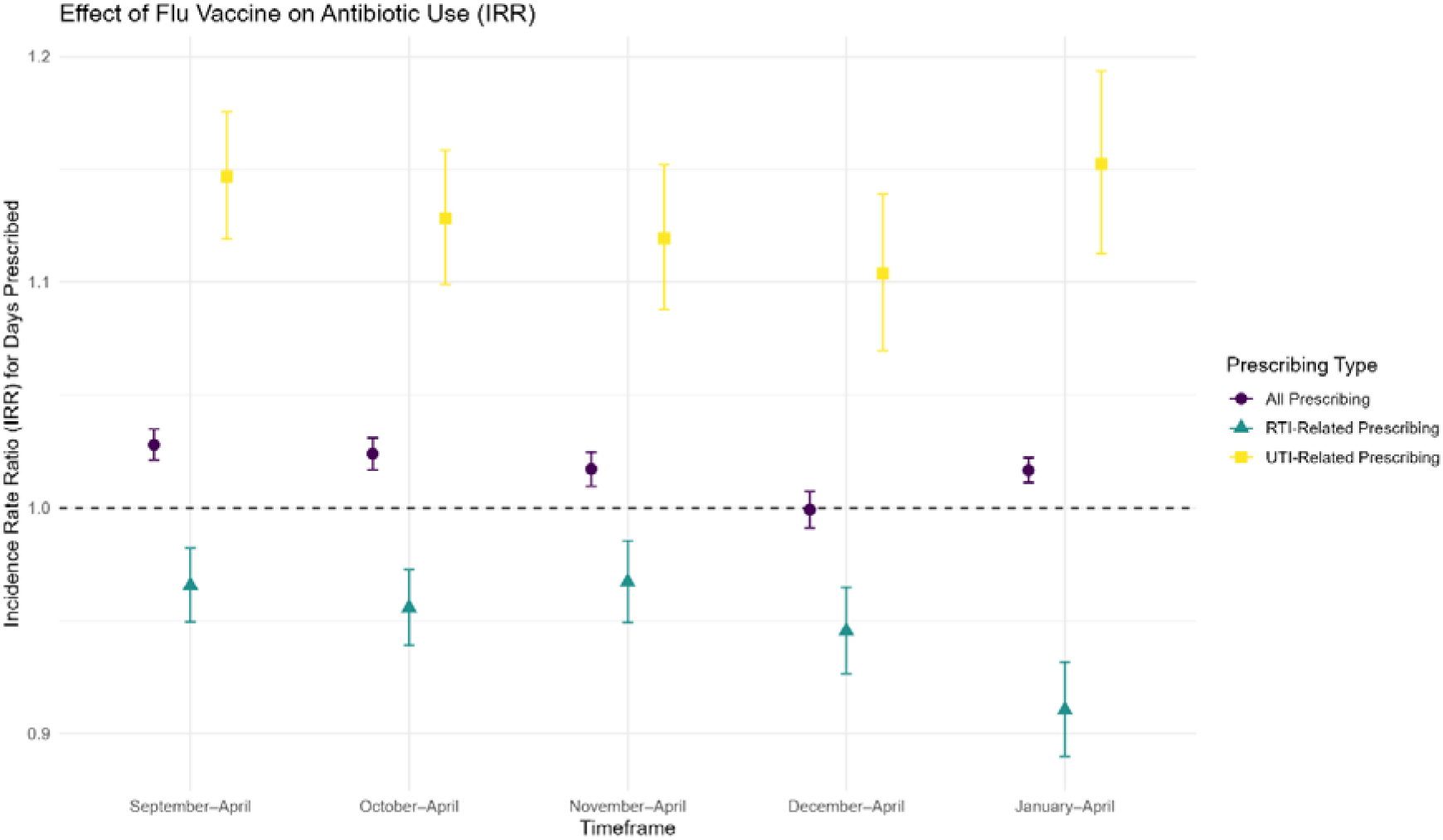
Incidence Rate Ratio (IRR) for the effect of influenza vaccination on antibiotic prescribing days by prescribing type (All, RTI-related, and UTI-related) across different timeframes (September-April, October-April, November-April, December-April, and January-April, each maintaining April as the fixed end date to narrow progressively toward peak influenza circulation). Shaded areas indicate the 95% confidence intervals.

In the multivariable models (Tables 2 and 3) other associations with prescribing included herpes zoster vaccination, with a statistically significant association with reduced number of days prescribed for any antibiotics by 6 percent; pneumococcal vaccine shows reduced RTI-related prescribing by between 9 and 16 percent; every interaction with a clinician is associated with an increase in prescribing days by 2 percent; and a higher combined sum comorbidity score shows an increase in prescribing by 4 percent per unit.

**Table 2:** Multivariable self-controlled case series conditional Poisson regression analysis for predictors of number of days prescribed antibiotics (September-April time window).

| Variable | Incidence Rate Ratio for all antibiotic prescriptions (95% CI) | Incidence Rate Ratio for RTI-related antibiotic prescriptions (95% CI) | Incidence Rate Ratio for UTI-related antibiotic prescriptions (95% CI) |
| --- | --- | --- | --- |
| **Influenza vaccine** | 1.03 (1.02–1.03) | 0.97 (0.95–0.98) | 1.15 (1.12–1.18) |
| **Influenza season** |  |  |  |
| 2011/12 | 1 | 1 | 1 |
| 2012/13 | 1.04 (1.04–1.05) | 1.06 (1.05–1.07) | 1.05 (1.03–1.08) |
| 2013/14 | 1.08 (1.07–1.08) | 0.99 (0.98–1.01) | 1.07 (1.05–1.09) |
| 2014/15 | 1.15 (1.14–1.15) | 1.13 (1.12–1.15) | 1.16 (1.14–1.18) |
| 2015/16 | 1.20 (1.20–1.21) | 1.08 (1.06–1.09) | 1.40 (1.37–1.43) |
| 2016/17 | 1.22 (1.21–1.23) | 1.17 (1.16–1.19) | 1.59 (1.55–1.62) |
| 2017/18 | 1.24 (1.24–1.25) | 1.22 (1.20–1.24) | 1.61 (1.57–1.64) |
| 2018/19 | 1.17 (1.16–1.18) | 0.97 (0.95–0.99) | 1.65 (1.62–1.69) |
| **Herpes zoster vaccine** | 0.94 (0.93–0.94) | 0.87 (0.86–0.88) | 0.85 (0.83–0.86) |
| **Pneumococcal vaccine** | 1.01 (1.00–1.02) | 0.91 (0.89–0.93) | 1.05 (1.01–1.09) |
| **Patient-clinical staff Interaction count** | 1.02 (1.02–1.02) | 1.03 (1.03–1.03) | 1.04 (1.04–1.04) |
| **Combined sum comorbidity score** | 1.04 (1.04–1.04) | 1.03 (1.03–1.04) | 1.05 (1.05–1.06) |
| **Out-of-season prescribing** | 1.01 (1.01–1.01) | 1.01 (1.01–1.01) | 1.01 (1.00–1.01) |
| **Centred age\*influenza vaccine** | 1.00 (1.00–1.00) | 1.00 (1.00–1.01) | 1.00 (0.99–1.00) |

**Table 3:** Multivariable self-controlled case series conditional Poisson regression analysis for predictors of number of days prescribed antibiotics (January-April time window).

| Variable | Incidence Rate Ratio for all antibiotic prescriptions (95% CI) | Incidence Rate Ratio for RTI-related antibiotic prescriptions (95% CI) | Incidence Rate Ratio for UTI-related antibiotic prescriptions (95% CI) |
| --- | --- | --- | --- |
| **Influenza vaccine** | 1.02 (1.01–1.02) | 0.91 (0.89–0.93) | 1.15 (1.11–1.19) |
| **Influenza season** |  |  |  |
| 2011/12 | 1 | 1 | 1 |
| 2012/13 | 1.02 (1.02–1.02) | 1.07 (1.05–1.08) | 1.04 (1.01–1.07) |
| 2013/14 | 1.06 (1.06–1.07) | 0.94 (0.92–0.96) | 1.02 (0.99–1.05) |
| 2014/15 | 1.12 (1.11–1.12) | 1.12 (1.09–1.14) | 1.18 (1.15–1.22) |
| 2015/16 | 1.20 (1.19–1.21) | 1.11 (1.09–1.14) | 1.42 (1.37–1.46) |
| 2016/17 | 1.22 (1.22–1.23) | 1.18 (1.15–1.20) | 1.59 (1.55–1.64) |
| 2017/18 | 1.24 (1.24–1.25) | 1.28 (1.25–1.31) | 1.63 (1.58–1.69) |
| 2018/19 | 1.20 (1.20–1.21) | 0.97 (0.95–1.00) | 1.66 (1.61–1.72) |
| **Herpes zoster vaccine** | 0.94 (0.94–0.95) | 0.83 (0.81–0.84) | 0.85 (0.82–0.87) |
| **Pneumococcal vaccine** | 1.00 (0.99–1.00) | 0.84 (0.82–0.87) | 1.03 (0.98–1.08) |
| **Patient-clinical staff**<br>**Interaction count** | 1.02 (1.02–1.02) | 1.03 (1.03–1.03) | 1.04 (1.04–1.04) |
| **Combined sum comorbidity**<br>**score** | 1.04 (1.03–1.04) | 1.03 (1.02–1.03) | 1.04 (1.03–1.05) |
| **Out-of-season prescribing** | 1.01 (1.01–1.01) | 1.01 (1.00–1.01) | 1.00 (1.00–1.00) |
| **Centred age\*flu vaccine** | 1.00 (1.00–1.00) | 1.01 (1.01–1.01) | 0.99 (0.99–1.00) |

Sensitivity analyses produced similar results to the main model estimates. These included models excluding herpes zoster vaccination covariate, unadjusted models including only the influenza vaccination indicator, models restricting the vaccinated period to 14 days post-vaccination or excluding individuals vaccinated after 30 November, and a complete-case analysis excluding patient-seasons with imputed prescribing duration (Supplementary Tables S1-S3, S6-S8 and Figures S2-S3). As a negative control, off-season antibiotic prescribing (May-August of the prior year) showed a modest positive association with subsequent-season influenza vaccination (IRR 1.08, 95% CI 1.07-1.09) that was consistent across timeframes, suggesting some residual confounding by unmeasured health-seeking behaviour not fully captured by the SCCS’s within-person design (Supplementary Table S5). Sex-stratified analysis indicated that this association differed by sex: the IRR was below 1 in men (0.83, 95% CI 0.79-0.88) and above 1 in women (1.25, 95% CI 1.22-1.29) for September-April, a pattern consistent across all narrower timeframes (Table S9).

## Discussion

The findings indicate that influenza vaccination is associated with a meaningful reduction in RTI-related antibiotic prescribing among individuals aged over 65. In contrast, no reduction was observed in overall antibiotic prescribing; and an increase in UTI-associated prescribing was evident during vaccinated seasons. These results suggest that while influenza vaccination may help reduce RTI-linked antibiotic use, the role of influenza vaccine in reducing antibiotic prescribing in general is less clear.

To our knowledge, this is the first study to evaluate the association between influenza vaccination and antibiotic prescribing in individuals aged over 65 using the self-controlled case series (SCCS) method. While cohort studies, such as one suggesting reduced amoxicillin prescribing in older adults (24), have addressed this topic, they are limited by confounding by indication, as healthier individuals are more likely to be vaccinated and have lower baseline antibiotic use. The SCCS method overcomes this limitation by controlling for time-invariant individual factors, making it in principle well-suited for evaluating seasonal vaccine effectiveness.

To further assess the impact of influenza vaccination on RTI-associated antibiotic prescribing, we investigated progressively shorter seasonal timeframes for analysis, increasing the probability of influenza-associated respiratory infections as the period of observation concentrated on the months of peak influenza transmission in the UK (25). We anticipated that the vaccine’s effect would be most pronounced during the height of influenza season.

The incidence rate ratio (IRR) for RTI-related prescribing decreased progressively with these shorter timeframes, consistent with the vaccine’s impact related to prevention of influenza and adverse influenza outcomes. These results highlight the importance of observation studies targeting peak influenza season, when evaluating non-specific endpoints, such as RTIs and prescribing, to ensure the outcomes align with the anticipated mechanism of action.

RTIs are the major cause of antibiotic prescribing in primary care and despite the finding of a reduction in RTI-prescribing with influenza vaccination, there was no change in overall prescribing and the surprising finding of an increase in UTI-associated prescribing. A biological mechanism to connect influenza vaccine with increasing UTI risk is lacking, however other potential explanations exist. We considered UTI prescriptions during the influenza season but predating vaccination as a possible explanation. However, a post-hoc analysis treating vaccination as a time-varying covariate within season did not fundamentally alter the IRRs. A more likely explanation is that increased GP attendances for potential urinary issues increases the probability of vaccination. We attempted to adjust for this by including number of GP interactions, comorbidity score, and out-of-season antibiotic prescribing as covariates in the model to account for variation in healthcare-seeking behaviour and underlying prescribing risk; however, this may not fully capture in-season patterns. Although this was a SCCS, it is expected that individuals’ behaviour will change year on year based on their state of health and the more GP attendances the more opportunities there will be to receive influenza vaccine. This may be particularly important in those individuals who do not regularly take influenza vaccines. We have attempted to adjust for this pre-season GP interaction, but this may not fully account for the in-season activity. Thus, UTI prescribing may be more causal to influenza vaccination than consequential.

Further exploration of this finding through a sex-stratified sensitivity analysis (Table S9) revealed markedly divergent effect estimates: the IRR for UTI-related prescribing was below 1 in men (0.83, 95% CI 0.79–0.88, September–April) but above 1 in women (1.25, 95% CI 1.22–1.29), a pattern that persisted across all narrower seasonal timeframes. This divergence is consistent with the explanation above rather than a genuine biological effect of vaccination on UTI risk, since the underlying drivers of a UTI diagnosis differ substantially by sex. In women, UTI-related presentations are common and frequently made on clinical grounds with limited microbiological confirmation (26) and may therefore be more sensitive to incidental contact with primary care, such as a vaccination visit, than to a change in true infection risk. In men, UTI-related prescribing is more often secondary to co-existing genitourinary conditions, such as prostatic pathology (27), which are not obviously triggered or unmasked by a vaccination consultation. If the pooled UTI association reflected a true consequence of vaccination, a broadly similar direction of effect would be expected in both sexes; the opposing estimates instead suggest that behavioural and diagnostic factors specific to women are principally responsible for the pooled increase. This interpretation remains exploratory, as the SCCS design compares vaccinated and unvaccinated seasons within the same individual and cannot fully distinguish confounding by healthcare contact from a true, sex-differential biological effect. Directly relating the timing of UTI-related prescriptions to the vaccination date within each season, separately by sex, would help establish whether the vaccination visit itself is the proximate trigger for UTI diagnosis in women, and is a natural next step for this work.

Pneumococcal and live shingles vaccination were incorporated as covariates in our models to account for the potential benefits of pneumococcal vaccine on reducing bacterial respiratory infections and live shingles vaccine as another indicator of healthcare seeking behaviour related to vaccines. A limitation of this proxy is that the live shingles vaccine used during the study period (Zostavax) is contraindicated in immunocompromised individuals; healthcare-seeking behaviour in this subgroup may therefore not be captured by this covariate. Notably these covariates were associated with a reduced risk of RTI prescribing and, in the case of shingles vaccine, overall prescribing. A post-hoc sensitivity analysis excluding herpes zoster vaccination did not significantly alter the IRRs for influenza vaccination and prescribing, suggesting that inclusion of this covariate did not materially affect the main findings. Whilst there is biological and epidemiological plausibility to these findings, we did not design the study to specifically investigate the impact of these vaccines on prescribing. We chose *a-priori* to use a SCCS because of the seasonal administration of influenza vaccinations; both pneumococcal vaccination and live shingles vaccination are not seasonally administered. Therefore, conclusions taken from these findings should be treated cautiously, as other confounding factors cannot be excluded. Furthermore, a systematic review indicated limited evidence of pneumococcal vaccination impacting on prescribing in adults, and there is no published evidence of live shingles vaccine impact on prescribing to date.

Although the reduction in RTI-related antibiotic prescribing with influenza vaccination was modest at an individual level, these findings suggest that sustained high uptake of influenza vaccination could translate into meaningful reductions in antibiotic use at the population level, contributing to efforts to reduce antimicrobial resistance. It should be noted that this study measured overall antibiotic use rather than inappropriate or unnecessary prescribing specifically. In the case of the latter this needs stewardship interventions (e.g. diagnostics, treatment guidelines) beyond vaccination.

### Strengths and limitations

This study has several key strengths that enhance the robustness and reliability of its findings. The SCCS methodology is a particularly valuable approach, as it inherently accounts for confounding by individual-level factors that remain constant over time. This is especially relevant in vaccine studies, where factors such as baseline health status and healthcare-seeking behaviour can significantly influence outcomes. By comparing vaccinated and unvaccinated timeframes within the same individual, this method mitigates much of the bias associated with traditional cohort designs.

The study’s large dataset, encompassing eight influenza seasons, is another strength. This extended period provides substantial statistical power and allows for detailed examination of seasonal trends and vaccine effectiveness across multiple flu seasons. Additionally, the inclusion of a partially vaccinated cohort offers unique insights that would not be achievable in analyses restricted to consistently vaccinated or unvaccinated populations. For example, vaccination coverage within the study cohort (58-69%) was lower than the nationally reported estimate of 72% (12), as the SCCS design requires exclusion of individuals vaccinated in every season. Baseline characteristics of the partially vaccinated and always-vaccinated cohorts are compared in Table S4. Furthermore, the observed reduction in RTI-linked antibiotic prescribing during vaccinated seasons is broadly consistent with the level of protection against medically attended influenza reported in UK primary care for the same population and study period (13), supporting the biological plausibility of the estimated effect even within a partially vaccinated cohort.

The use of prescribing days as the outcome measure provides a more detailed quantification of antibiotic exposure compared to the number of prescriptions alone. By capturing the duration and intensity of antibiotic courses, this measure offers a clearer evaluation of the potential changes in antibiotic use associated with influenza vaccination.

Despite its strengths, this study has limitations. In this study, we opted not to include time-varying covariates in our analysis. Similar studies using the SCCS method have measured prescribing outcomes as time-varying covariates to capture sufficient detail across two influenza seasons (28). Unlike these studies, our analysis utilized data from eight seasons.

This longer timeframe provided sufficient granularity without the need for time-varying covariates, which avoided potential overfitting to seasonal fluctuations. By treating all covariates as non-time-varying (within season), we were able to remove seasonality effects from the analysis while simplifying model structure. This approach ensured that our findings reflected the relationship between influenza vaccination and antibiotic prescribing without overfitting to seasonal trends. Nonetheless, we conducted a supplementary post-hoc analysis treating vaccination as a time-varying covariate within season, which confirmed that this modelling choice did not materially alter the estimated IRRs.

While this approach ensured model simplicity and consistency across seasons, it also meant that vaccination status was assigned for the entire September-April period in any season where an individual received the vaccine, regardless of their actual vaccination date within that period. This may have introduced exposure misclassification. As a result, prescribing events that occurred before vaccination may have been incorrectly attributed to the vaccinated period, potentially biasing effect estimates. We also formally tested whether prior-season antibiotic prescribing predicted subsequent vaccination uptake within individuals – a potential source of cross-season confounding if healthcare-seeking behaviour simultaneously drives both outcomes. A logistic regression of vaccination status on prior out-of-season prescribing days, adjusted for influenza season, produced an odds ratio of 1.004 (95% CI: 1.004-1.005), indicating a statistically significant but small association. This suggests that while a trend exists for individuals with higher prior prescribing to be more likely to vaccinate in the following season, the magnitude of this relationship is unlikely to materially bias the primary IRR estimates.

Another limitation relates to the length of follow-up. We did not explicitly account for increasing frailty among ageing members of the cohort, which could influence healthcare-seeking behaviour and prescribing patterns over time. While this is unlikely to have had a major impact when comparing a few consecutive influenza seasons, it may become more important across longer periods of follow-up as a greater proportion of individuals develop frailty. The direction of such an effect is difficult to predict, as frailty may either increase the likelihood of both vaccination and prescribing or reduce healthcare contact altogether. Future work should consider incorporating measures of frailty when examining long-term vaccine effects in elderly populations.

It is also possible that trends unrelated to influenza vaccination, such as antimicrobial stewardship initiatives implemented over the study period, contributed to the observed decline in prescribing. The within-person SCCS comparison, combined with the season-level covariate included in our model, mitigates confounding from such trends to some extent, but cannot fully exclude a residual stewardship effect if its impact varied differentially between an individual’s vaccinated and unvaccinated seasons.

A limitation inherent to the SCCS design is that this analysis cannot include interactions for demographic measures such as sex, ethnicity, and Index of Multiple Deprivation (IMD).

Further, although the SCCS design strengthens causal inference by controlling for fixed individual-level confounders, the findings should be interpreted as associations rather than definitive causal effects. It is also worth noting that every vaccine is treated as an individual effect rather than a cumulative effect of sequential annual vaccination.

We did not attempt to translate the observed IRR into a population-level estimate of prescriptions avoided. The SCCS design estimates a within-person relative effect among individuals who were vaccinated in some but not all seasons; this population differs systematically from those who are consistently vaccinated or unvaccinated, who were excluded from the analysis by design. Extrapolating the observed IRR to the wider 65-and-over population would therefore require assumptions about generalizability that our design cannot support, and risks conveying spurious precision for a modest relative effect. We instead present the IRR itself, alongside its confidence interval, as the primary measure of effect. Another limitation is that the study does not adjust for the effectiveness of the seasonal vaccine in each season or the timing of the vaccine with respect to the peak of the flu season. This is because in the SCCS framework, vaccine effectiveness is defined at the seasonal level and is therefore perfectly colinear with influenza season covariate included in our analysis. However, season-to-season variation in influenza severity was still accounted for indirectly through the categorical influenza season term included in all models. We were therefore unable to formally test whether the vaccine-prescribing association varied with the severity of a given influenza season; this would be a useful extension for datasets with finer-grained surveillance linkage.

### Further work

The findings of this study highlight several directions for future research. The elevated incidence of UTI-linked prescribing during vaccinated periods likely reflects residual confounding as discussed above. This underscores the value of refining exposure definitions in SCCS analyses, particularly for outcomes not directly related to the vaccine’s mechanism of action. Future studies could explore alternative methodological approaches to better align outcome timing with vaccine administration, helping to clarify true vaccine effects versus methodologically driven patterns.

We also need to economically contextualize the findings, but a large part of this will depend on a better understanding of prescribing density and AMR generation which is currently imprecise.

Reproducibility is also a key consideration. Conducting similar analyses in other datasets or populations would help validate the findings and assess their generalisability. Populations with different age distributions, healthcare access patterns, or prescribing behaviours could reveal whether the effects observed in this study are consistent across diverse contexts.

## Conclusion

We found a significant reduction in antibiotic prescribing in vaccinated older adults during periods of active influenza circulation. These findings may support the conclusion that influenza immunisation can contribute to effectively reduce the burden of antibiotic prescribing in primary care, especially during peak influenza seasons. While we also found evidence that pneumococcal and live shingles vaccination may reduce RTI-related prescribing, these findings should be investigated in robustly designed intervention specific studies. Nevertheless, our study advocates for the increased uptake and use of immunisation against respiratory pathogens in older populations.

## Footnotes

The authors have completed the STROBE checklist, which is included as supplementary material.

## Acknowledgements

The authors acknowledge the providers of data for this study and thank the general practices involved. We would like to thank Heather Whitaker for her guidance and feedback on this study.

## Data availability

The datasets used in this study were extracted from Clinical Practice Research Datalink (CPRD) following CPRD approval of the study protocol (reference 21_000457: available at https://cprd.com/protocol/) and through a multi-study license and data sharing agreement between the University of Liverpool and CPRD. The Medicines and Healthcare products Regulatory Agency and National Institute for Health and Care Research sponsor CPRD. The authors are not authorised to share the datasets and are obliged to destroy the datasets according to the data-sharing agreement between the University of Liverpool and CPRD.

## Contributors

DH and NF contributed equally. DH, NF conceptualised the study. DS acquired the data. DS, TS, and DH carried out the statistical analyses. NF, DH, MH, and DS contributed to the design of methodology and models. DH, MH, NF contributed to supervision. DS and TS carried out visualisation/data presentation. TS wrote the original draft of the manuscript. DH, NF, MH, TS, DS, RV and VD contributed to writing, reviewing, and editing the manuscript. DH and NF are the guarantors and accept full responsibility for study conduct, had access to the data and controlled the decision to publish. The corresponding author attests that all listed authors meet authorship criteria.

## Ethics and Consent to participate

All data in this study were anonymised. Clinical Practice Research Datalink’s Research Data Governance (RDG) granted approval (reference 21_000457). Rolling ethical approval is obtained for CPRD from the Health Research Authority (reference number 05/MRE04/87).

