## Supplementary Tables/Figures for "The Impact of Influenza Vaccination on Antibiotic Prescribing in Older Adults: A Self-Controlled Case Series Analysis"

**Supplementary Material for: The Impact of Influenza Vaccination on Antibiotic Prescribing in Older Adults**

### Additional cohort descriptives

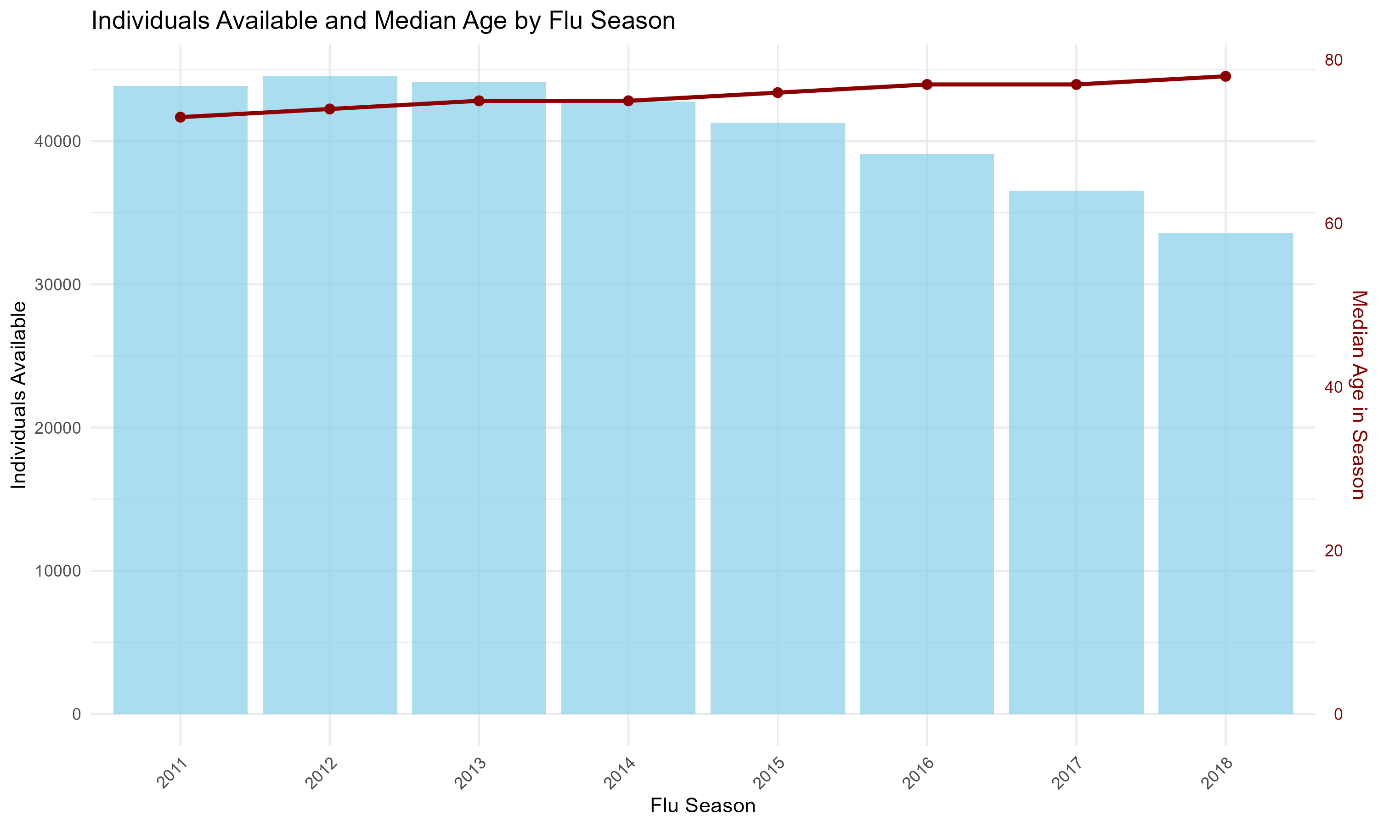

**Figure S1: Number of individuals available and median age by influenza season (2011/12-2018/19). Bars (left axis) show the count of individuals contributing to each season; the dark red line (right axis) shows median age in season.**

### Sensitivity analysis: removing herpes zoster vaccination variable

Table S1: Multivariable self-controlled case series conditional Poisson regression analysis for predictors of number of days prescribed antibiotics, excluding herpes zoster vaccine covariate (September–April time window).

| **Variable** | **Incidence Rate Ratio for all antibiotic prescriptions (95% CI)** | **Incidence Rate Ratio for RTI-related antibiotic prescriptions (95% CI)** | **Incidence Rate Ratio for UTI-related antibiotic prescriptions (95% CI)** |
| --- | --- | --- | --- |
| **Influenza vaccine** | 1.03 (1.02–1.03) | 0.96 (0.95–0.98) | 1.14 (1.11–1.17) |
| **Influenza season** |  |  |  |
| 2011/12 | 1 | 1 | 1 |
| 2012/13 | 1.04 (1.04–1.05) | 1.06 (1.05–1.07) | 1.05 (1.03–1.07) |
| 2013/14 | 1.07 (1.07–1.08) | 0.99 (0.97–1.00) | 1.06 (1.04–1.08) |
| 2014/15 | 1.14 (1.13–1.14) | 1.11 (1.10–1.13) | 1.13 (1.11–1.16) |
| 2015/16 | 1.19 (1.18–1.19) | 1.05 (1.03–1.06) | 1.35 (1.33–1.38) |
| 2016/17 | 1.20 (1.19–1.21) | 1.13 (1.11–1.15) | 1.52 (1.49–1.55) |
| 2017/18 | 1.22 (1.21–1.23) | 1.17 (1.15–1.19) | 1.53 (1.49–1.56) |
| 2018/19 | 1.15 (1.14–1.15) | 0.92 (0.91–0.94) | 1.56 (1.52–1.60) |
| **Pneumococcal vaccine** | 1.00 (0.99–1.01) | 0.89 (0.87–0.91) | 1.02 (0.99–1.06) |
| **Patient-clinical staff Interaction count** | 1.02 (1.02–1.02) | 1.03 (1.03–1.03) | 1.04 (1.04–1.04) |
| **Combined sum comorbidity score** | 1.04 (1.04–1.04) | 1.03 (1.03–1.04) | 1.05 (1.05–1.06) |
| **Out-of-season prescribing** | 1.01 (1.01–1.01) | 1.01 (1.01–1.01) | 1.01 (1.00–1.01) |
| **Centred age*influenza vaccine** | 1.00 (1.00–1.00) | 1.00 (1.00–1.01) | 1.00 (0.99–1.00) |

Table S2: Multivariable self-controlled case series conditional Poisson regression analysis for predictors of number of days prescribed antibiotics, excluding herpes zoster vaccine covariate (January–April time window).

| **Variable** | **Incidence Rate Ratio for all antibiotic prescriptions (95% CI)** | **Incidence Rate Ratio for RTI-related antibiotic prescriptions (95% CI)** | **Incidence Rate Ratio for UTI-related antibiotic prescriptions (95% CI)** |
| --- | --- | --- | --- |
| **Influenza vaccine** | 1.01 (1.01–1.02) | 0.90 (0.88–0.93) | 1.14 (1.10–1.18) |
| **Influenza season** |  |  |  |
| 2011/12 | 1 | 1 | 1 |
| 2012/13 | 1.02 (1.02–1.02) | 1.07 (1.05–1.08) | 1.04 (1.01–1.07) |
| 2013/14 | 1.06 (1.06–1.06) | 0.93 (0.91–0.95) | 1.01 (0.98–1.04) |
| 2014/15 | 1.11 (1.11–1.12) | 1.09 (1.07–1.11) | 1.16 (1.12–1.19) |
| 2015/16 | 1.19 (1.18–1.19) | 1.07 (1.05–1.09) | 1.37 (1.33–1.41) |
| 2016/17 | 1.20 (1.20–1.21) | 1.12 (1.10–1.14) | 1.53 (1.48–1.57) |
| 2017/18 | 1.22 (1.21–1.23) | 1.20 (1.17–1.22) | 1.55 (1.50–1.60) |
| 2018/19 | 1.18 (1.17–1.18) | 0.90 (0.88–0.93) | 1.57 (1.52–1.62) |
| **Pneumococcal vaccine** | 0.99 (0.98–1.00) | 0.82 (0.80–0.85) | 1.00 (0.96–1.05) |
| **Patient-clinical staff Interaction count** | 1.02 (1.02–1.02) | 1.03 (1.03–1.03) | 1.04 (1.04–1.04) |
| **Combined sum comorbidity score** | 1.04 (1.03–1.04) | 1.03 (1.02–1.03) | 1.05 (1.04–1.06) |
| **Out-of-season prescribing** | 1.01 (1.01–1.01) | 1.01 (1.01–1.01) | 1.00 (1.00–1.00) |
| **Centred age*influenza vaccine** | 1.00 (1.00–1.00) | 1.01 (1.01–1.01) | 0.99 (0.99–1.00) |

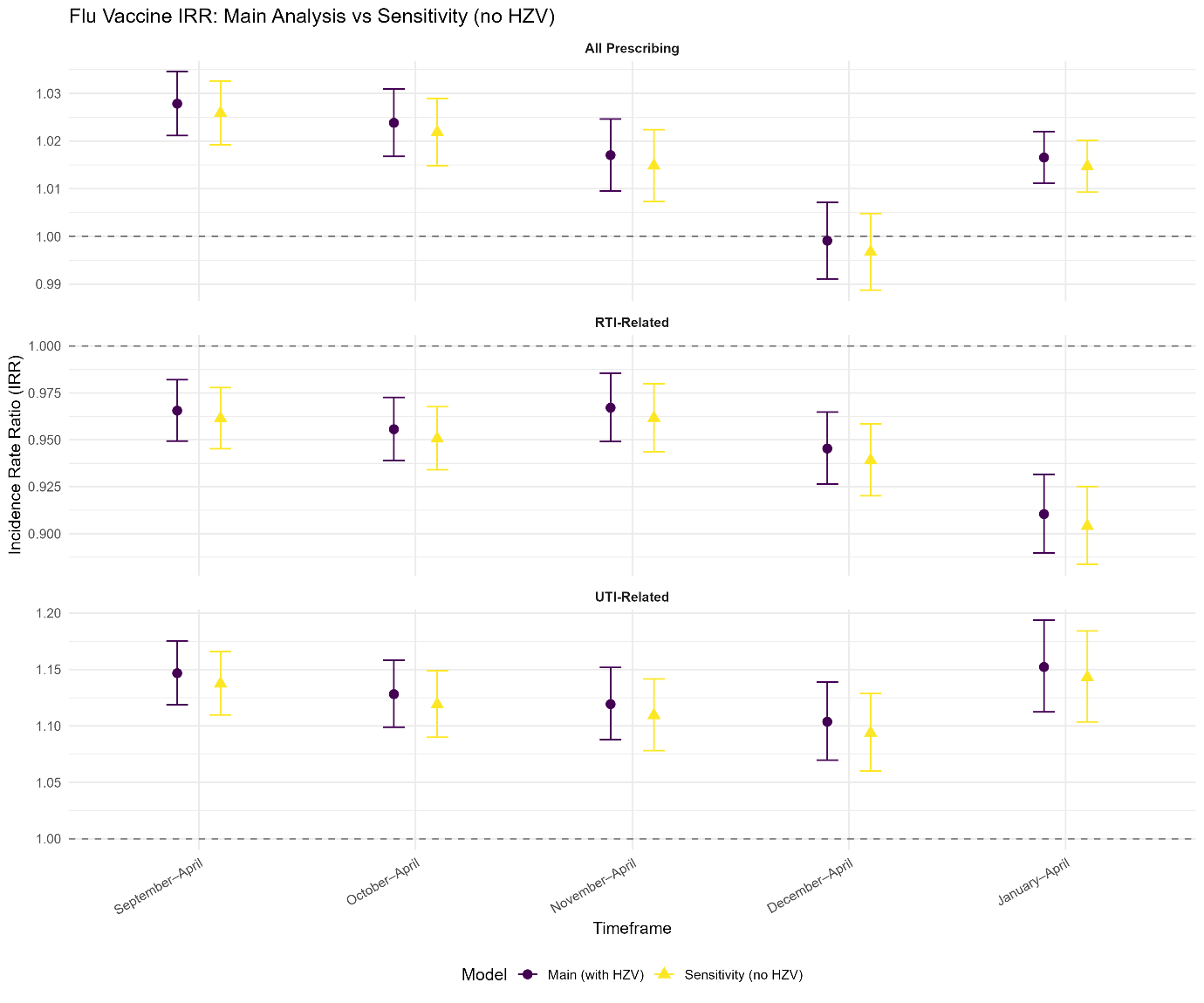

**Figure S2: Incidence Rate Ratio (IRR) for the effect of influenza vaccination on antibiotic prescribing days by prescribing type (All, RTI-related, and UTI-related) across different seasonal timeframes (September–April, October–April, November–April, December–April, and January–April, each maintaining April as the fixed end date to narrow progressively toward peak influenza circulation): main analysis (with herpes zoster vaccine covariate) versus sensitivity analysis (without herpes-zoster vaccine covariate). Points show IRR estimates; error bars indicate Wald 95% confidence intervals.**

### Unadjusted analysis: including only influenza vaccination as an exposure

**Table S3: Univariable self-controlled case series conditional Poisson regression analysis estimating the association between seasonal influenza vaccination and number of days prescribed antibiotics (unadjusted models).**

| **Variable** | **Incidence Rate Ratio for all antibiotic prescriptions (95% CI)** | **Incidence Rate Ratio for RTI-related antibiotic prescriptions (95% CI)** | **Incidence Rate Ratio for UTI-related antibiotic prescriptions (95% CI)** |
| --- | --- | --- | --- |
| **September-April** | 1.08 (1.08–1.09) | 1.07 (1.06–1.08) | 1.11 (1.10–1.12) |
| **October-April** | 1.08 (1.08–1.08) | 1.07 (1.06–1.08) | 1.10 (1.09–1.12) |
| **November-April** | 1.07 (1.07–1.08) | 1.07 (1.06–1.08) | 1.10 (1.08–1.11) |
| **December-April** | 1.06 (1.05–1.06) | 1.06 (1.05–1.07) | 1.10 (1.08–1.11) |
| **January-April** | 1.07 (1.06–1.07) | 1.03 (1.02–1.07) | 1.08 (1.06–1.10) |

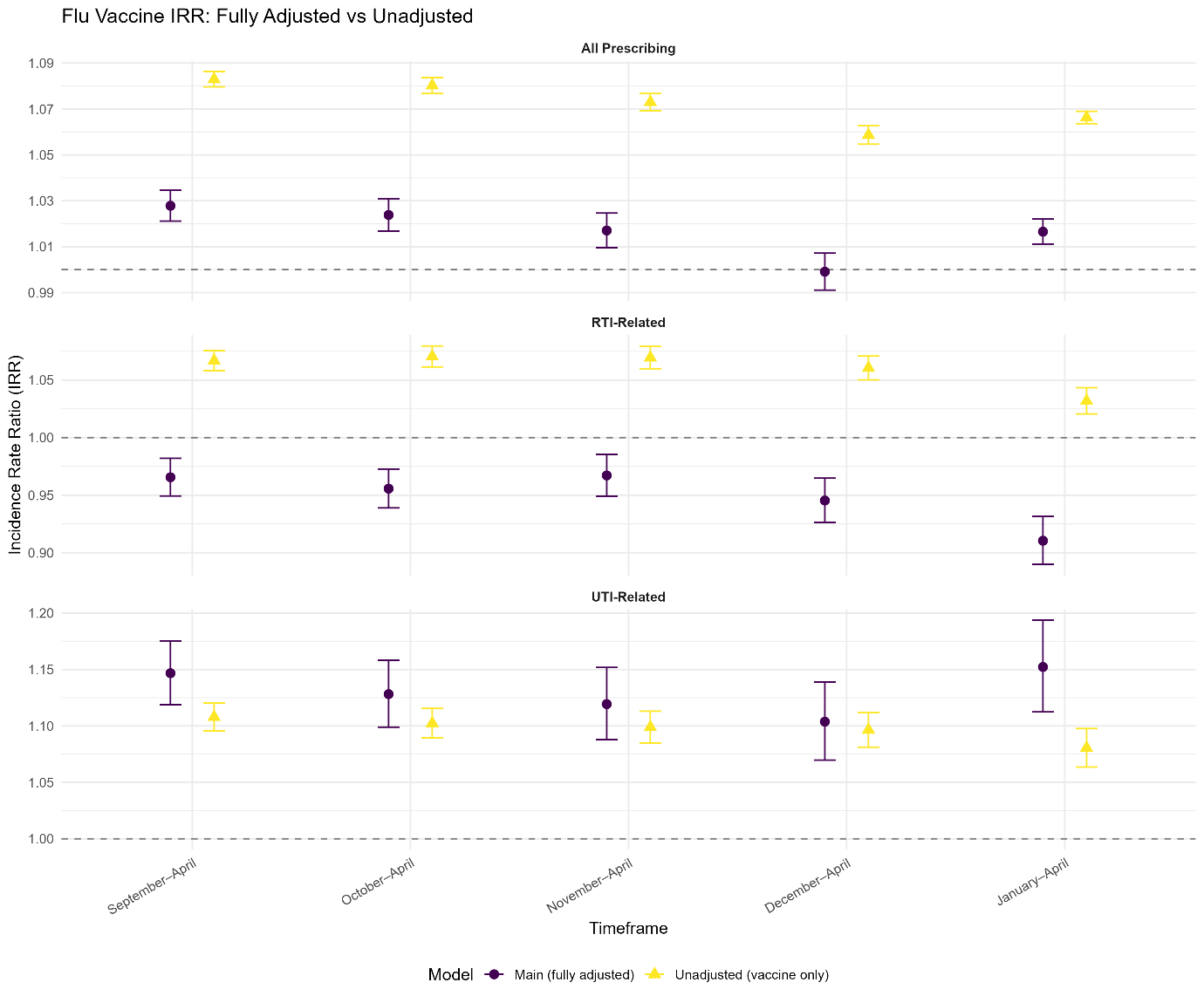

**Figure S3: Incidence Rate Ratio (IRR) for the effect of influenza vaccination on antibiotic prescribing days by prescribing type (All, RTI-related, and UTI-related) across different seasonal timeframes (September–April, October–April, November–April, December–April, and January–April, each maintaining April as the fixed end date to narrow progressively toward peak influenza circulation): fully adjusted main analysis versus unadjusted model (influenza vaccine indicator only). Points show IRR estimates; error bars indicate Wald 95% confidence intervals.**

### Additional cohort descriptives: Partially vs fully vaccinated comparison

**Table S4: Baseline characteristics of the analysed partially vaccinated cohort compared with excluded fully vaccinated individuals. Median [IQR] reported for continuous variables; proportions reported for categorical variables.**

| **Group** | **Partially vaccinated (analysed cohort)** | **Always vaccinated (excluded)** |
| --- | --- | --- |
| **N** | 48329 | 136752 |
| **Median age [IQR]** | 76.5 [71.5, 82.5] | 78.5 [73.5, 84.0] |
| **% Female** | 57.9% | 55.1% |
| **% IMD most deprived** | 14.8% | 13.1% |
| **% IMD least deprived** | 24.5% | 26.2% |
| **Comorbidity score, median [IQR]** | 0.0 [0.0, 1.0] | 0.5 [0.0, 1.0] |

### Sensitivity analysis: Off-season antibiotic prescribing as a negative control outcome

**Table S5: Influenza vaccine IRR (95% CI) for off-season antibiotic prescribing days across seasonal timeframes. Off-season prescribing days (May-August of the prior year) were modelled as the outcome using the same covariate structure as the primary analysis, excluding the out-of-season prescribing covariate.**

| **Timeframe** | **Influenza vaccine IRR (95% CI)** |
| --- | --- |
| **September-April** | 1.08 (1.07–1.09) |
| **October-April** | 1.08 (1.07–1.09) |
| **November-April** | 1.08 (1.07–1.09) |
| **December-April** | 1.08 (1.07–1.09) |
| **January-April** | 1.08 (1.07–1.09) |

### Sensitivity analysis: restricting vaccinated period to at least 14 days post-vaccination date

**Table S6: Influenza vaccine IRR (95% CI) restricting the vaccinated period to at least 14 days post-vaccination date, across seasonal timeframes and antibiotic prescribing outcomes. Individuals that did not meet the partially vaccinated criterion were excluded (n=703 individuals removed).**

| **Timeframe** | **All-cause IRR (95% CI)** | **RTI-linked IRR (95% CI)** | **UTI-linked IRR (95% CI)** |
| --- | --- | --- | --- |
| **September-April** | 1.03 (1.02–1.04) | 0.98 (0.97–1.00) | 1.14 (1.11–1.17) |
| **October-April** | 1.03 (1.02–1.04) | 0.98 (0.96–0.99) | 1.13 (1.10–1.16) |
| **November-April** | 1.02 (1.02–1.03) | 0.99 (0.97–1.01) | 1.12 (1.09–1.16) |
| **December-April** | 1.01 (1.00–1.02) | 0.97 (0.95–0.99) | 1.11 (1.08–1.15) |
| **January-April** | 1.02 (1.02–1.03) | 0.94 (0.91–0.96) | 1.17 (1.13–1.21) |

### Sensitivity analysis: excluding individuals vaccinated after 30 November

**Table S7: Influenza vaccine IRR (95% CI) excluding individuals vaccinated after 30 November (over 91 days after 1 September), across seasonal timeframes and antibiotic prescribing outcomes. Individuals that did not meet the partially vaccinated criterion were excluded (n=2,117 individuals removed).**

| Timeframe | **All-cause IRR (95% CI)** | **RTI-linked IRR (95% CI)** | **UTI-linked IRR (95% CI)** |
| --- | --- | --- | --- |
| September-April | 1.03 (1.02–1.04) | 0.93 (0.91–0.94) | 1.12 (1.09–1.15) |
| October-April | 1.02 (1.02–1.03) | 0.92 (0.90–0.93) | 1.11 (1.08–1.14) |
| November-April | 1.01 (1.00–1.02) | 0.94 (0.92–0.95) | 1.09 (1.06–1.12) |
| December-April | 0.99 (0.99–1.00) | 0.93 (0.92–0.95) | 1.05 (1.02–1.08) |
| January-April | 1.02 (1.02–1.03) | 0.93 (0.91–0.95) | 1.10 (1.06–1.13) |

### Sensitivity analysis: complete case analysis

**Table S8: Influenza vaccine IRR (95% CI) for September-April antibiotic prescribing outcomes, main analysis vs complete case (patient-seasons requiring imputed prescription duration excluded), 1159 individuals were removed relative to the main analysis cohort either through the complete case restriction itself or through reapplication of the partially-vaccinated criteria on the reduced data.**

| **Timeframe** | **Main analysis IRR (95% CI)** | **Complete case IRR (95% CI)** |
| --- | --- | --- |
| **All Prescribing** | 1.03 (1.02–1.03) | 1.02 (1.02–1.03) |
| **RTI-related** | 0.97 (0.95–0.98) | 0.98 (0.96–1.00) |
| **UTI-related** | 1.15 (1.12–1.18) | 1.11 (1.08–1.14) |

### Sensitivity analysis: UTI-related prescribing outcome stratified by sex

**Table S9: UTI-related antibiotic use: influenza vaccine IRR (95% CI) by sex and seasonal timeframe**

| **Timeframe** | **Male IRR (95% CI)** | **Female IRR (95% CI)** |
| --- | --- | --- |
| **September–April** | 0.83 (0.79–0.88) | 1.25 (1.22–1.29) |
| **October–April** | 0.77 (0.73–0.81) | 1.26 (1.22–1.30) |
| **November–April** | 0.81 (0.76–0.85) | 1.24 (1.20–1.28) |
| **December–April** | 0.84 (0.79–0.89) | 1.20 (1.16–1.24) |
| **January–April** | 0.81 (0.75–0.87) | 1.30 (1.25–1.35) |
